# Design of a Behavioural Risk Monitoring Tool (RutiSafeNet) in an Acute Inpatient Mental Health Unit with Open-Doors Policy: A Qualitative Study

**DOI:** 10.1101/2025.10.01.25337049

**Authors:** Laura Miró Mezquita, Anna Moreno Orea, Joan de Pablo Rabasso, Maria Isabel Martínez Segura, Marina Delgado Marí, Sandra Castro Anso, María Jiménez Murcia, Ana Ibañez Caparros, Tatiana Bustos Cardona, Jorge Cuevas-Esteban

## Abstract

**Introduction:** Open-door policies for acute inpatient mental health units (AIMHU) have shown promising results in reducing coercive measures, but concerns remain among patients and staff regarding increased workload, constant surveillance, and potential safety risks associated with these policies.

**Objectives:** This study aims to develop a monitoring tool to facilitate safety management in AIMHUs by monitoring behavioural risks and nursing workload.

**Methods:** This study employed a qualitative approach using the content analysis method. Data were collected through three in-depth interviews and two focus groups involving staff (n=19) from the AIMHU of the hospital.

**Results:** 34 items were identified to define behavioural risks related to self-harm and suicide, aggressiveness, and absconding, alongside factors affecting nursing workload. These items were categorised into three levels of risk: low, moderate, and high.

**Conclusions:** The result is a risk monitoring tool based on observational items for daily use in AIMHU that can help to reduce the uncertainty about risk management, especially in open-door units. This innovative tool enables individualized patient monitoring, provides insights into the unit’s status, prioritizes conflict prevention, and upholds the human rights of patients.

## 1. INTRODUCTION

Human rights have emerged as a central focus in the evaluation of mental health policies and strategies (Herrman and Swartz, 2007; Patel et al., 2007) with The World Health Organization (WHO) urging all nations to safeguard the rights of individuals hospitalised with mental disorders (Scobie et al., 2006). The structure of psychiatric inpatient wards, significantly influences treatment outcomes and patient experience (Krückl et al., 2023). While traditional closed-door policies are designed to enhance safety and security, they are often perceived by both patients (Haglund & Von Essen, 2005) and staff (Haglund et al., 2006) as restrictive and non-therapeutic environments. These perceptions may contribute to emotional distress and hinder recovery (Johnson et al., 2022). The primary justification for closed-door policies is to ensure safety, particularly for preventing suicide and aggressive behaviour, and reducing the risk of absconding incidents (Haglund et al., 2007; Scobie et al., 2006). However, the employment of coercive measures and the lack of freedom of movement undermines patients’ fundamental rights, thus posing significant challenges in hospital management and nursing practice (Kennedy et al., 2019). In response to this issue, new hospitalisation models have been developed that aim to create a safe environment while minimizing the need for such coercive measures (Gooding et al., 2020).

Risk factors associated with various conflictive behaviours in mental health units have been extensively investigated. Powell et al. (Powell et al., 2000) conducted a study focusing on suicide risk factors in hospitalised patients, identifying several indicators including suicidal ideation, prior self-harm attempts, recent bereavement, presence of delusions, chronic mental illness, and family history of suicide. In assessing the risk of imminent aggression on a day-to-day basis, the Dynamic Appraisal of Situational Aggression – Inpatient Version (DASA-IV) (Ogloff & Daffern, 2006) scale was developed for adult psychiatric inpatients. Moreover, tools like the Waypoint Elopement Risk Scales (WERS) (Marshall & Usinger, 2016) have been developed to assess the risk of absconding in psychiatric inpatients, taking into account both clinical history, such as abscondment or previous attempts, wandering, substance abuse, self-harm, and harm to others; and day-to-day events, such as absconding threats, substance use, life stressors, current risk to self-harm and medication adherence.

It is also deemed essential to monitor the nursing workload required by each patient according to their clinical needs, while establishing risk monitoring to ensure safety. The nursing workload contributes to establishing the global situation in relation to risk management and deciding whether the doors can be opened (Petrucci et al., 2014). In a scoping review published in 2018 (Sousa & Seabra, 2018) on the evaluation of nursing workload in adult psychiatric inpatient units, four studies were identified (Fanneran et al., 2015; Gerolamo, 2009; Petrucci et al., 2014; Twigg & Duffield, 2009). Petrucci et al. (Petrucci et al., 2014) devised a nursing care complexity indicator tool predicated on the personal mental health history extracted from patients’ medical records upon admission. This tool comprises 88 items correlated with Gordon’s 11 functional health patterns (Gordon, 1982). The remaining studies investigate how to assess workload to better distribute workforce planning. The systematic review concludes that few instruments exist to evaluate or measure nursing workload in adult patients hospitalised in mental health units, and those available, lack uniformity. (Rosen et al., 2018; Van Den Oetelaar et al., 2021). None of these instruments enable the continuous detection of patient-associated workload changes throughout the entire hospitalisation process.

In the context of mental health hospitalisation, especially in open-door policies, it is crucial to assess patients’ behavioural risks and nursing workload to ensure the safety of both patients and staff. To the best of our knowledge, there are no instruments that, based on observation, objectively evaluate both behavioural risks and nursing workload simultaneously and throughout the entire hospitalization process for each patient. Therefore, the aim of the study is to design an inpatient monitoring tool based on objective items for categorising patients’ behavioural risks and nursing workload to ensure the safety of patients and staff while maintaining an open-door policy.

## 2. METHODOLOGY

### 2.1. Study Design

The aim of this study is to design a monitoring tool for inpatients that allows for the establishment of categories and quantification of the level of behavioural risk and nursing workload to facilitate global security management and offer an objective measure for the decision to open doors. To address the objective of this study, a qualitative investigation was undertaken through individual interviews and focus groups during the first quarter of 2023. Before initiating the qualitative study, the interviewer, that is the first researcher, conducted an observational analysis of the unit to manage interview time effectively, prioritize topics based on their importance, and prevent participant dispersion.

An expert panel, composed of psychiatrists, a nurse manager and senior mental health nurses, initially defined the two main categories, behavioural risks and nursing workload. Based on the literature, it was also considered relevant to subdivide the behavioural risk category into subcategories: self-harm and suicide, aggression, and absconding. The tool is designed to monitor each patient in both main categories, behavioural risk and nursing workload, throughout the entire hospital stay. To classify the levels of behavioural risk and nursing workload, the expert committee defined three levels as a starting point: low, moderate, and high, associated with values of 0, 1, and 2, respectively. Each patient receives a score between 0 and 2 for each main category, giving equal importance to both categories. The sum of the scores of all patients enables the therapeutic team to numerically assess the environmental safety.

### 2.2. Setting

The present study was conducted at the Acute Inpatient Mental Health Unit of Germans Trias University Hospital (GTUH), Badalona, Barcelona, Spain, a general multispecialty hospital affiliated to the University Autonoma of Barcelona. At the time of the study, GTUH catchment area was of 350.530 adults distributed across four community mental health areas in the North Barcelona health district, with a mean of 313 adult discharges per year. The acute inpatient psychiatric unit at GTUH consists of one ward with 20 beds and are staffed by multidisciplinary teams of senior and junior psychiatrists, nurses, psychologists, and social workers. The unit provides multidisciplinary acute specialist clinical care for patients admitted with serious mental illnesses, in the form of pharmacotherapy, psychotherapy, social work and occupational care, and a recovery program. Since 2019, the unit has progressively implemented the Safewards care model. Safewards is a set of ten interventions designed to enhance safety by preventing conflict and containment (Bowers, 2014). Electro-convulsive therapy is available when the study was carried out. Most of the patients (90-95%) are admitted from the emergency room. They usually suffer from severe primary psychiatric disorders with acute disturbances or risk of self-harm. The remaining of patients are referred from community mental health providers. The ward has open-door policies, meaning that door was open during daytime, and at night-time the door was closed. These policies have been conducted from October 2021.

### 2.3. Rigour

This research was conducted according to the guidelines in the Standards for Reporting Qualitative Research (SRQR) (O’Brien et al., 2014). To establish credibility, independent coding of the data was done by two researchers, followed by a collaborative consensus on emerging codes. Transferability was addressed by providing detailed descriptions of the AIMHU setting and the open-door policy context, allowing readers to assess the applicability of findings to their own environments. To ensure dependability, we have clearly outlined the data collection and analysis processes, including the interviews and focus groups methodologies. The study adhered to ethical guidelines, receiving approval from the local research ethics committee (PI-23-143).

### 2.4. Participants

The inclusion criteria for participants were as follows: (1) be working in the hospital ‘s acute mental health unit, and (2) having experience, at least six months, working on Open-doors policies. The participants were selected using the purposive method. The individual interviews included 3 participants (66% female) from different areas of work, and they were contacted face-to-face. For the focus groups, a total of 29 participants were contacted by email and 16 agreed to take part in the study. Informed consent was obtained from all subjects involved in the study.

Two focus groups were conducted, with 8 participants per session. The participants included 8 nurses, 5 nursing assistants, 1 psychiatrist, 1 psychologist and 1 social worker. Of the participants, 81.25% were females, while the rest were males. The focus groups were held at the hospital facilities. The sociodemographic data for all the participants is shown in Table 1.

**Table 1.** Descriptive statistics of participants in the qualitative study.

|  | mean (years) | SD (years) |
| --- | --- | --- |
| Age | 36.61 | 10.83 |
| Work experience | 11.33 | 8.98 |
| Experience at the hospital | 8.09 | 8.68 |

### 2.5. In-depth semi-structured interviews

The data were collected through face-to-face semi-structured interviews conducted in a designated room within the AIMHU of the hospital. Only the researcher and the interviewee were present in the room. Prior to the interviews, an interview guide was developed by the expert panel.

Each interview was structured into two sections according to the categories: behavioural risks and nursing workload. Behavioural risk section was subdivided in the three different subcategories. Each section started with the following question: “Do you think it is appropriate to differentiate it into low, moderate and high level?”. It was followed by an open-ended query depending on the subtheme addressed: “What observations would you employ as criteria to delineate whether a patient is categorised as low, moderate, or high risk?”.

These initial prompts were succeeded by further exploratory inquiries for each behavioural risk, such as: “What behavioural characteristics are exhibited by patients at risk of aggression?” “What common traits do patients who attempt to abscond share?”.

To objectively assess nursing workload, the inquiries included the same initial questions for the levels of categorisation and initial items. Moreover, workload disparities between voluntary and involuntary inpatients were discussed and identifying physical care needs that require increased staff attention was a crucial issue. The influence of the mental status in the nursing workload was also explored.

The duration of the interviews averaged approximately 45 minutes for each participant. They were audio-recorded using two distinct devices, with participants providing verbal consent prior to commencement. Subsequently, the interviews were meticulously transcribed. Initial line-by-line coding was performed according to the content analysis method and descriptive codes were identified by two researchers independently. These “codes” are referred to as items throughout the article and, subsequently, in the developed tool. In accordance with the methodology of qualitative studies, the term “categories” will refer to themes, and “subcategories” will denote subthemes. Categories were defined in advance, that is, behavioural risks and nursing workload, and subcategories for behavioural risks were validated.

### 2.6. Focus groups

Two focus groups were conducted in a hospital training room, guided by two researchers (LMM and MIMS). Based on insights extracted from the analysis of interview data, a comprehensive guide was developed to direct the focus groups toward relevant topics. No additional individuals were present in the interview room aside from the participants and the researchers. During the discussions, items and the corresponding levels extracted from individual interviews were first introduced and deliberated upon. After that, other situations were exposed while researchers diligently documented field notes and identify new items as necessary. It was considered highly relevant to collect information on the appropriate duration of alert maintenance for each item. Each focus group extended over a duration of 1.5 hours, and data saturation was achieved in both instances. As in the interviews, the recordings were made with two different devices and transcribed after the session. The analysis of the information mirrored the content analysis methodology used in the interviews. After the interviews and focus groups, coding was compared and categorised according to similarities and differences. Finally, it was decided to convene the expert panel to refine, consolidate, and validate the items for each level.

## 3. RESULTS

The result of this study is a tool for monitoring acute patients in mental health units during admission, which will be called RutiSafeNet.

### 3.1. Behavioural risks

First, the debate revolved around which types of situations should be included in the tool. Collectively, they agreed that, since the tool aims to monitor changes during hospitalization, it should focus more on clinical presentation and individual behaviours rather than clinical history and intrinsic patient information.

“I believe it’s more about the clinical presentation than the patient.” (P9)

Regarding self-harm and suicide risk, all participants unanimously agreed that constant verbalization of suicidal ideation poses a high risk, while a punctual verbalization of self-harm wishes represented a moderate risk. Any type of self-injury or medication overdose was also agreed upon to merit a high score. “And a high risk, well, that it comes from a well-structured suicidal thought, or because it has been carried out or is manifesting that it is going to do it already without telling you how. Also, that directly during the shift it is evidenced that self-injury or medication overdose has been performed.” (P1)

Picking up dangerous objects such as sharp objects, lighters, and belts was identified as posing a high risk of aggressiveness to both themselves and others. Additionally, there was discussion about the severity of verbal threats, with the minimum score being moderate risk.

“When in doubt, I would always mark it as red because you never know what could happen, but the type of verbal threat should be considered.” (P2)

Regarding absconding risk, hypervigilance with access points and requests for personal documentation were mentioned as indicators of considerable risk. Previous absconding attempts were considered high risk in involuntary inpatients.

“If they come in very decompensated and have a lot of history of absconding, it’s more likely that they will leave than if they come in for a change in treatment or because they are awaiting another device.” (P6)

### 4.2. Nursing workload

The definition of nursing workload score revolved around the cognitive status of patients and their demands.

“Very high is a person who is all the time in the control room, asking for our attention, that you have to be with him during the shift, that is very invasive, very reiterative.” (P5)

“Medium level, would be a depression for example that needs us to be there doing reconductions.” (P1)

Patients requiring moderate demands such as assistance with hygiene or meal companionship were deemed to impose a moderate workload. Patients with a high degree of dependence and those requiring constant redirection were identified as posing the highest burden, warranting a score of two.

Finally, Table 2 presents the 34 items and their corresponding score and duration for categorising risks associated with self-harm and suicide, aggressiveness, and absconding, as well as the nursing workload attributed to patients.

**Table 2.** Items, score and duration for self-harm and suicide, aggressiveness and absconding risk and nursing workload extracted from the complete qualitative study.

|  | Items | Score (level) | Duration |
| --- | --- | --- | --- |
| **Behavioural risks** |  |  |  |
| *Self-harm and suicide risk* |  |  |  |
|  | Verbalization of suicidal ideation | 2 | Three shifts |
|  | Verbalization of passive death wishes | 1 | One shift |
|  | Moderate/severe self-harm incidents | 2 | Three shifts |
|  | Mild self-harm during shift | 1 | One shift |
|  | Suicidal attempts | 2 | Three shifts |
|  | Hiding an object with potential for self/hetero harm | 2 | Three shifts |
|  | Stockpiling medication | 2 | Three shifts |
|  | Suspicion of suicidal risk expressed by the family | 1 | Three shifts |
|  | Hiding in blind spots of the camera | 1 | One shift |
|  | Psychomotor pre-agitation | 2 | Three shifts |
| *Aggressiveness risk* |  |  |  |
|  | Aggressive behaviour towards people | 2 | Three shifts |
|  | Aggressive behaviour towards objects | 2 | Three shifts |
|  | Verbal aggressiveness | 1 | One shift |
|  | Threatening behaviour | 1 | Three shifts |
|  | Recently unrestrained patient | 2 | Three shifts |
| *Absconding risk* |  |  |  |
|  | Explicit verbal manifestation | 2 | Three shifts |
|  | Hypervigilance with access points | 2 | Three shifts |
|  | Constant demands for discharge | 1 | One shift |
|  | Refusal to take medication | 1 | One shift |
|  | Acute cognitive change | 1 | One shift |
|  | Previous absconding attempts in involuntarily admitted patients | 2 | Entire admission |
|  | Moderate/severe cognitive impairment with ambulation | 1 | Entire admission |
|  | Involuntary admission patient | 1 | Until voluntary admission |
| Nursing workload |  |  |  |
|  | Demanding patient | 1 | One shift |
|  | Patient dependent for activities of daily living | 2 | One shift |
|  | Patient requiring assistance with hygiene | 1 | One shift |
|  | Patient in environmental restraint | 2 | One shift |
|  | Isolation due to contagious condition | 2 | One shift |
|  | Patient with eating disorder | 1 | One shift |
|  | Patient in mechanical restraint | 2 | One shift |
|  | Requires constant accompaniment | 2 | One shift |
|  | Requires intermittent accompaniment | 1 | One shift |
|  | Requires nursing care for acute somatic pathology | 1 | One shift |
|  | Reversal of oppositional behaviour | 1 | One shift |

1: moderate risk/workload
2: high risk/workload

## 4. DISCUSSION

This study presents the development of a clinical risk score tool for managing patient safety in mental health hospitalisation. The tool has been successfully designed to provide a rapid and efficient assessment of the therapeutic climate and environmental safety through a total score within AIMHUs. The tool is based on 34 items, differentiated into two main categories, behavioural risks and nursing workload and specifically assesses self-harm and suicide, aggression and absconding risk. Categories and subcategories were considered appropriated in the study, aligning with the risk division defined by the National Health Service (2006) (Scobie et al., 2006) and Haglund et al. (Haglund et al., 2007). The distinction into three levels— low, moderate, and high— was also regarded as suitable for each category and subcategory, and the majority consider it acceptable.

Regarding self-harm and suicide risk, it was concluded that verbalization of suicidal ideation, moderate or severe self-harm, and suicide attempts address high risk, coinciding with the conclusions drawn by Powel et al. (Powell et al., 2000). In the present study it has been reported that the severity of suicidal ideation can be differentiated into two risk levels depending on the severity. Passive death wishes and non-suicidal self-injury have been considered of moderate risk. Furthermore, events that are not intrinsic to the patient have not been taken into consideration, such as previous family history of suicide or recent bereavement because the tool is designed to be based on observational items.

Items of the aggressive category that score as high risk are aggressive behaviour towards people, towards objects and recently unrestrained patient. Verbal aggressiveness and threatening behaviour are considered medium risk. The DASA-IV (Ogloff & Daffern, 2006) scale item ‘Verbal threats’ is directly related to verbal aggression, but the four remaining items extracted from this qualitative study do not have an equivalence in the DASA-IV scale.

In respect to absconding risk subcategory items, explicit verbal expression was considered a high risk, as also defined in the WERS scale (Marshall & Usinger, 2016). Hypervigilance with access points, which has been concluded to pose a high risk of absconding, is not included in the WERS scale. On the other hand, within this subcategory, it has been established that previous absconding attempts only represent a risk in patients with involuntary admissions, whereas the WERS scale considers them regardless of the type of admission. F Although the WERS scale does not address this issue, Georgieva et al. (Georgieva et al., 2012) reported that it had high predictive power for seclusion and the use of coercive measures. The medium risk item ‘refusal to take medication’ is also included in the WERS scale, while ‘acute cognitive change’ can be reflected in the WERS item ‘behaviours.’ The remaining items from this qualitative study that indicate a medium risk of absconding—namely, constant demands for discharge, moderate to severe cognitive impairment with ambulation, and involuntary admission—do not have equivalents in the aforementioned scale.

Finally, the nursing workload category includes items related to patient care, excluding those associated with administrative tasks. In this study, different levels of workload (low, moderate, and high) have been distinguished, while in the tool developed by Petrucci et al. (Petrucci et al., 2014), all items receive the same score. One of the major differences between this tool and the scale developed by Petrucci is that the latter only considers the nursing workload at the time of admission, whereas ours assesses it during each shift. This enables the acquisition of current information regarding patient needs and the associated workload. In the “health management” category of Petrucci’s tool, acute somatic pathology care and management of patients with oppositional behaviours are included; in the “Activity/exercise” category, items related to dependency on basic activities of daily living are considered; and in “Nutritional,” patients irregularly consuming food or liquids are scored, although it may not be related directly to an eating disorder. The oppositional behaviours are also included in the DASA-IV scale. Some situations covered in the 88-item tool are also found in the one extracted from this qualitative study; however, they are not included in the nursing workload category. Items related to supervision (constant and intermittent accompaniment and demanding patient) are not considered in the Petrucci’s tool. The remaining items, patients in mechanical or environmental restraint, or with a contagious condition, have been classified in different levels in the nursing workload category.

This designed tool could facilitate the reduction of negative perceptions and emotions related to staff burnout, safety, and excessive responsibility, and it would help alleviate the uncertainty associated with security. This tool directly addresses the safety concerns raised by staff and patients regarding open-door policies, as highlighted in several qualitative studies (Indregard et al., 2024; Kalagi et al., 2018; Muir-Cochrane et al., 2012; Sollied et al., 2023). Since the tool provides numerical values, algorithms can be developed to generate a score that reflects the overall climate of the unit. The aggregation of both parameters offers insight into environmental safety and allows for the establishment of a cut-off point to guide decisions regarding the opening of doors.

One of the strengths of the current study is that it was based on two focus groups, and 3 in-depth interviews with the collaboration of a wide variety of mental health professionals. Employing qualitative techniques like semi-structured interviews enables participants to elaborate on their answers, leading to a comprehensive data gathering process that captures depth and intricate details beyond what a quantitative method can achieve. To the best of our knowledge, it is the only study that simultaneously considers the behavioural risks of hospitalized patients and the workload they generate for the nursing staff. This innovative tool manages to include factors not only considered in the past patients’ history and enhances existing scales. It is worth noting that RutiSafeNet, may be modifiable and adaptable to other types of patients and units, such as locked door wards. Moreover, due to the way the information in the tool is structured, organised into categories and subcategories, and the concise nature of the items, it is a user-friendly tool that requires minimal prior training.

### Limitations

This study has some limitations. One of the primary limitations of this study is that the developed instrument has not yet been validated through quantitative studies or comparisons with established scales. While the tool has been designed based on qualitative evidence and expert consensus, its validity and reliability still need to be evaluated in future research. Specifically, further studies are required to assess the instrument’s internal consistency, temporal stability, and its relationship with other measures of risk and workload in mental health units. Without these analyses, the application of the instrument should be considered preliminary, and its clinical use should be approached with caution until empirical data support its effectiveness and accuracy. Secondly, it’s possible that there might be a bias in opinions among the professionals since all of them are members of the same acute unit staff. Since open-door policy had been recently implemented, the participants’ experience might not be enough. Thirdly, in the nursing workload category, non-patient related activities, such as training other nurses and organisational and administrative work, have not been considered, but their impact on the overall level of nursing workload could be analysed. More evidence and the conduct of new studies in other AIMHUs with different policies would be necessary to verify the congruence of the items.

## 5. CONCLUSIONS

This study underscores the importance of structured risk assessment in AIMHUs, particularly in the context of open-door policies. By identifying and categorizing 34 key items related to behavioural risks and nursing workload, we have developed a practical monitoring tool that facilitates daily risk management. This tool provides a standardized approach to evaluating self-harm, aggressiveness, and absconding risks, supporting clinical decision-making and enhancing patient safety. The implementation of this monitoring tool enables individualized patient monitoring and provides a real-time overview of unit dynamics, contributing to conflict prevention. Further research is required to validate the scale, enhance its applicability across diverse healthcare settings, and assess its long-term impact on reducing coercive therapeutic measures. Nevertheless, the standardization of risk control and workload monitoring represents an advancement in the management of AIMHUs, with the potential for broader implementation at higher levels.

## Data Availability

All data produced in the present study are available upon reasonable request to the authors

## Acknowledgments

The authors would like to thank the participants of the qualitative study for giving their precious time in assisting the sessions.

